# Increased risk of infection with SARS-CoV-2 Omicron compared to Delta in vaccinated and previously infected individuals, the Netherlands, 22 November to 19 December 2021

**DOI:** 10.1101/2021.12.20.21268121

**Authors:** Dirk Eggink, Stijn P. Andeweg, Harry Vennema, Noortje van Maarseveen, Klaas Vermaas, Boris Vlaemynck, Raf Schepers, Arianne B. van Gageldonk-Lafeber, Susan van den Hof, Chantal B.E.M. Reusken, Mirjam J. Knol

**Author notes:** contributed equally (Dirk Eggink, Stijn P. Andeweg). contributed equally (Chantal B.E.M. Reusken, Mirjam J. Knol).

## Abstract

Infections by the Omicron SARS-CoV-2 variant are rapidly increasing worldwide. Among 70,983 infected individuals (age ≥ 12 years), we observed an increased risk of S-gene target failure, predictive of the Omicron variant, in fully vaccinated (odds ratio: 5.0; 95% confidence interval: 4.0-6.1) and previously infected individuals (OR: 4.9: 95% CI: 3.1-7.7) compared with infected naïve individuals. This suggests a substantial decrease in protection from vaccine- or infection-induced immunity against SARS-CoV-2 infections caused by the Omicron variant compared with the Delta variant.

---

On 26 November, 2021, the WHO declared Omicron SARS-CoV-2 (Nextclade 21K, Pangolineage B.1.1.529) a variant of concern^1^. Since then, the Omicron variant has been rapidly spreading across the world^2^. The United Kingdom and Denmark currently report a very rapid increase of infections caused by Omicron, showing the higher transmissibility of this variant^3^. ECDC expects Omicron to be the dominant variant in Europe within a few weeks^4^. Omicron displays an unprecedented number of alterations in the Spike protein, including around 30 amino acid substitutions, 3 deletions and 1 insertion^5–7^. These alterations raise concerns about the protection evoked by current SARS-CoV-2 vaccines against the Omicron variant.

Here we investigate variant distribution of Delta and Omicron among SARS-CoV-2 positive individuals who were either naive, fully vaccinated or had a previous infection to investigate possible escape from vaccine- or infection-induced immunity by Omicron compared to Delta.

## Data

We used data from two large diagnostic laboratories analyzing specimens from national community testing in the Netherlands that make use of the TaqPath COVID-19 RT-PCR Kit (ThermoFisher Scientific). Omicron possesses a deletion at amino acid positions 69 and 70 of the spike protein (Δ69–70) that has been associated with failure of the probe targeting the S-gene, while the Orf1ab and N probe result in a proper signal (S-gene target failure (SGTF), also referred to as S-drop-out). This failure of detection of the S-gene target in an otherwise positive PCR test has proven to be a highly specific proxy for Alpha in the past and Omicron in current days in a background of sporadic circulation of other Δ69–70 variants ^8–11^. However, with lower viral loads, the S-gene target tends to be the least sensitive of the three targets in the Taqpath system, which could result in incorrect allocation of a SGTF. To avoid erroneous calling of SGTF, a stringent threshold was used to identify likely Omicron infections. In-house titration showed decreased sensitivity for the S-gene target in case of viral loads that give a cycle threshold (Ct) value of around 32 for the ORF1ab and N targets. Therefore, only results with a Ct value of ≤ 30 for both targets were included for further analyses.

S-gene testing results were linked to vaccination status data from the national notification register (OSIRIS), which includes all positive SARS-CoV-2 cases in the Netherlands. If vaccination status was unknown from the notification register, we used self-reported vaccination status from the community testing register (CoronIT). Persons were defined as fully vaccinated when they received two doses of Comirnaty (BNT162b2, BioNTech/Pfizer), Spikevax (mRNA-1273, Moderna) or Vaxzevria (ChAdOx1, AstraZeneca) more than 14 days before the symptom onset date or one dose of Janssen vaccine more than 28 days before the symptom onset date. For cases without an onset date, sample date minus two days was used. Persons that did not receive any vaccine were defined as unvaccinated. Previous infection was defined as a positive PCR or antigen test result at least 8 weeks before the current positive test based on the notification and testing register. We excluded children <12 years of age because they were not yet eligible for vaccination in the study period, and there have been different testing policies in this age group over time influencing the likelihood of detecting a previous infection.

## Statistical analysis

Among infected individuals we compared the percentage of SGTF between unvaccinated cases without a known previous infection (naïve), fully vaccinated cases without a known previous infection, unvaccinated cases with a known previous infection. The number of fully vaccinated persons with a previous infection was very small and therefore excluded. We performed logistic regression to estimate the association between immune status and SGTF, adjusting for testing date (natural cubic spline with 3 knots), 10-year age group and sex. As travel history could be related to being infected with the Omicron variant and with being vaccinated, we performed an additional analysis, where we excluded cases with a history of travel outside the Netherlands in the 14 days before symptom onset.

## Immune status and SGTF

Between 22 November and 19 December 2021, 70,983 PCR positive samples with a Ct value <30 for ORF1ab and N-gene targets were analyzed for SGTF and included in the current analysis. SGTF was present in 931 (1.3%) cases; the percentage of SGTF increased rapidly from early December (Figure 1). Cases with SGTF were younger and more often had a travel history (Table 1). Of the SGTF cases, 789 (85%) were fully vaccinated compared to 62% of non-SGTF cases and 2.8% had a previous infection compared to 1.3% of the non-SGTF cases. Correspondingly, we found an adjusted odds ratio of 5.0 (95% CI: 4.0-6.1) for the association between full vaccination and SGTF. Similarly, an adjusted odds ratio of 4.9 (95% CI: 3.1-7.7) was found for previous infection. When excluding cases with any known travel history, the odds ratio remained similar for previous infection (OR 4.7; 95%: 3.0-7.6) and was lower for vaccination but still highly significant (OR 4.3, 95% CI: 3.5-5.3).

**Table 1.** Characteristics of SARS-CoV-2 cases of 12 years and older with S-gene detected and S-gene target failure (SGTF), n= 70983, the Netherlands, 22 November - 19 December 2021

|  | **S-gene detected, N (%)** | **S-gene target failure, N (%)** |
| --- | --- | --- |
| Total | 70052 | 931 |
| Immune status |  |  |
| Naïve | 25684 (36.7%) | 116 (12.5%) |
| Full vaccination | 43423 (62.0%) | 789 (84.7%) |
| Previous infection | 945 (1.3%) | 26 (2.8%) |
| Age group |  |  |
| 12-19 | 8490 (12.1%) | 81 (8.7%) |
| 20-29 | 9689 (13.8%) | 232 (24.9%) |
| 30-39 | 13016 (18.6%) | 154 (16.5%) |
| 40-49 | 12800 (18.3%) | 139 (14.9%) |
| 50-59 | 10359 (14.8%) | 178 (19.1%) |
| 60-69 | 8911 (12.7%) | 106 (11.4%) |
| 70-79 | 5291 (7.6%) | 40 (4.3%) |
| 80+ | 1496 (2.1%) | 1 (0.1%) |
| Sex |  |  |
| Male | 35273 (50.4%) | 507 (54.5%) |
| Female | 34779 (49.6%) | 424 (45.5%) |
| Travel history |  |  |
| Yes | 578 (0.8%) | 128 (13.7%) |
| No | 31929 (45.6%) | 293 (31.5%) |
| Unknown | 37545 (53.6%) | 510 (54.8%) |
| Median number of days btw onset and last vaccination (interquartile range) | 152 (53) | 154 (44) |
| Median number of days btw onset and previous infection (interquartile range) | 337 (140) | 266 (212) |

**Table 2.**
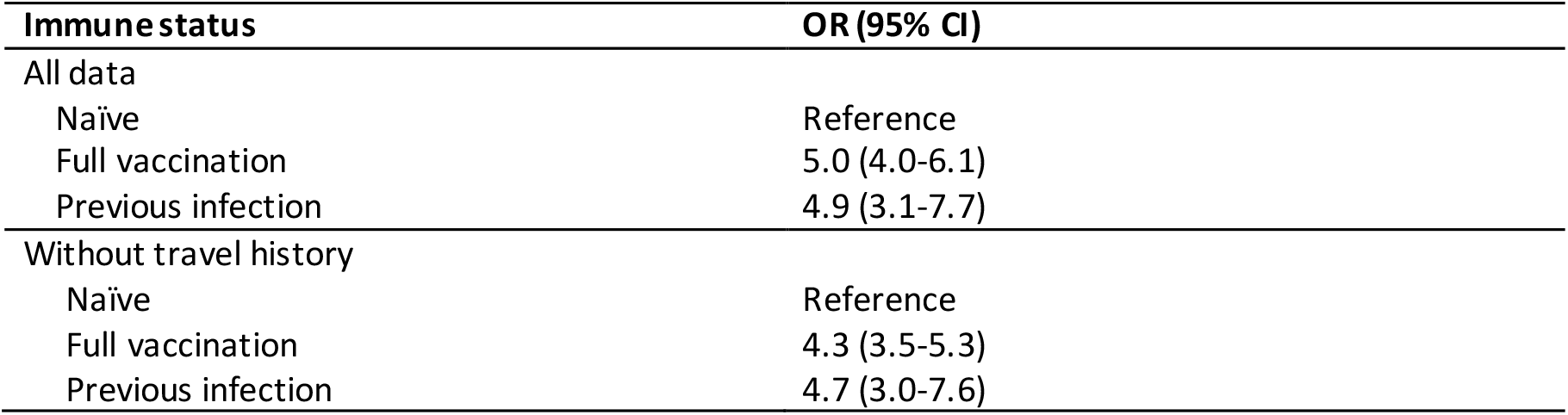
Odds ratios (ORs) and 95% confidence intervals (CIs) for the association between immune status and SGTF adjusted for day of sampling, sex and 10-year age group fitted to all data and only tests from cases without travel history

| **Immune status** | **OR (95% CI)** |
| --- | --- |
| All data |  |
| Naïve | Reference |
| Full vaccination | 5.0 (4.0-6.1) |
| Previous infection | 4.9 (3.1-7.7) |
| Without travel history |  |
| Naïve | Reference |
| Full vaccination | 4.3 (3.5-5.3) |
| Previous infection | 4.7 (3.0-7.6) |

**Figure 1.**
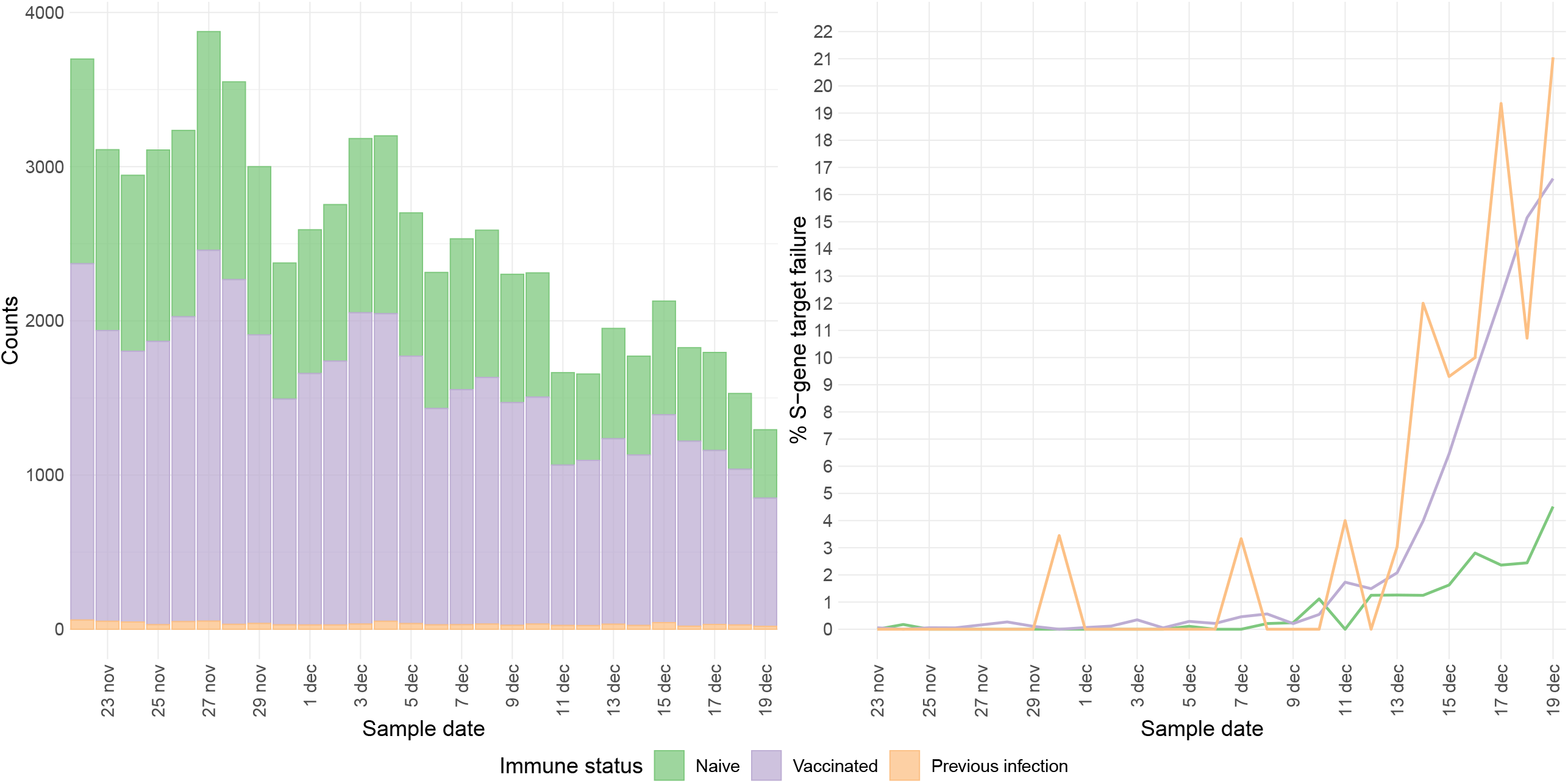
Number of included cases (left panel) and percentage of SGTF (right panel) by immune status, n= 70983, the Netherlands, 22 November - 19 December 2021

## Discussion

Our results suggest a large reduction in protection against SARS-CoV-2 infection with the Omicron variant compared to the Delta variant after full vaccination. This is in concordance with in vitro studies that showed a 30-40-fold reduction in neutralisation of the Omicron variant with convalescent sera or sera of fully vaccinated individuals ^5–7^. Booster vaccination appears to show a smaller reduction in neutralisation sensitivity^12^. The UK showed a substantial reduction in vaccine effectiveness against infection with the Omicron variant compared with Delta after primary vaccination^13,14^. After booster vaccination, the vaccine effectiveness against Omicron infection increased but was still lower than against Delta. Our data shows an OR of around 5 suggesting a significant reduction in vaccine effectiveness.

Despite small numbers, we showed that individuals with a previous infection also have a clear increased risk of infection with Omicron compared with naïve individuals, suggesting that previous infection with another SARS-CoV-2 variant provides low levels of protection against Omicron infection. This contrasts with what we observed in a previous analysis, where we did not find an increased risk of infection with the Delta, Beta and Gamma variants versus the Alpha variant in previously infected individuals^15^. For reinfections, we observed a shorter period between the two episodes for Omicron cases compared to Delta. Such difference between Omicron and Delta cases was not observed for time between last dose and infection for vaccinated cases. Our finding of reduced protection against reinfection with Omicron is in line with surveillance data from South Africa and England, showing increased risk of reinfections with the Omicron variant^16–18^.

Our study looked at infections based on community testing. It is still unclear what the influence of the Omicron variant is on the protection by vaccination or previous infection against severe COVID-19.

Denmark reports for now a lower crude proportion of hospitalization for Omicron than for Delta (based on 114 hospitalized Omicron cases), however this may be explained by younger age groups affected by Omicron or time lag for patients to be hospitalized^19^. In the UK, no clear change in severity was observed but the number of Omicron hospitalized cases was small (n=24)^11,14,18^. Even when disease severity by Omicron is lower compared with Delta, the higher transmissibility will probably lead to more hospital admissions when no additional measures are taken.

In our study we were not yet able to assess the effect of booster vaccination, as booster vaccination has only recently started in The Netherlands (18 November 2021). Our results did not change substantially when excluding cases with a travel history.

Our results suggest a large decrease in protection from vaccine- or infection-induced immunity against SARS-CoV-2 infections caused by the Omicron variant compared with the Delta variant. This emphasizes the urgent need for booster vaccination and will warrant implementing non-pharmaceutical interventions to prevent overflooding of hospital care if COVID-19 severity is not reduced to a great extent.

## Data Availability

Data produced in the present study are available upon reasonable request to the authors.

